# Estimating the effect of social inequalities in the mitigation of COVID-19 across communities in Santiago de Chile

**DOI:** 10.1101/2020.10.08.20204750

**Authors:** Nicolò Gozzi, Michele Tizzoni, Matteo Chinazzi, Leo Ferres, Alessandro Vespignani, Nicola Perra

**Affiliations:** Networks and Urban Systems Centre, University of Greenwich, London, UK; ISI Foundation, Turin, Italy; Laboratory for the Modeling of Biological and Socio-technical Systems, Northeastern University, Boston, MA USA; Data Science Institute, Universidad del Desarrollo, Santiago, Chile; Telefónica R&D, Santiago, Chile

## Abstract

We study the spatio-temporal spread of SARS-CoV-2 in Santiago de Chile using anonymized mobile phone data from 1.4 million users, 22% of the whole population in the area, characterizing the effects of non-pharmaceutical interventions (NPIs) on the epidemic dynamics. We integrate these data into a mechanistic epidemic model calibrated on surveillance data. As of August 1^st^ 2020, we estimate a detection rate of 102 cases per 1,000 infections (90% CI: [95 - 112 per 1,000]). We show that the introduction of a full lockdown on May 15^th^, 2020, while causing a modest additional decrease in mobility and contacts with respect to previous NPIs, was decisive in bringing the epidemic under control, highlighting the importance of a timely governmental response to COVID-19 outbreaks. We find that the impact of NPIs on individuals’ mobility correlates with the Human Development Index of *comunas* in the city. Indeed, more developed and wealthier areas became more isolated after government interventions and experienced a significantly lower burden of the pandemic. The heterogeneity of COVID-19 impact raises important issues in the implementation of NPIs and highlights the challenges that communities affected by systemic health and social inequalities face adapting their behaviors during an epidemic.

## 1 Introduction

As of September 1^st^, 2020, Chile has reported more than 400, 000 cases and 590 SARS-CoV-2 deaths per million, becoming one of the worst COVID-19 epidemic globally [1]. Officially, the first SARS-CoV-2 case in Chile was detected on March 3^*rd*^, 2020 [2]. Although other cases were rapidly confirmed all over the country, the urban area of the capital city, Santiago Metropolitan Region, quickly became the epicenter of the national epidemic. Indeed, as of September 1^st^, 2020, about 70% of the total cases in the nation have been reported in the *comunas* (i.e. municipalities) of Santiago, making it one of the largest urban COVID-19 outbreak in the world. The first set of non-pharmaceutical interventions (NPIs) were put in place in mid-March, when schools were closed, public gatherings were banned, and passengers traveling from high-risk countries were mandated to self-isolate for 14 days. However, the adopted measures were not able to contain the contagion: after a sharp increase in cases, a full lockdown was instituted to the whole area of Santiago on May 15^*th*^ [3].

In this work, we model the spatial and temporal spread of COVID-19 in 37 *comunas* of the urban area of Santiago. We aim to provide a data-driven characterization of the unfolding of the COVID-19 epidemic and an estimation of the impact of NPIs on its spreading. To this end, we study the reduction of mobility and contacts inferred from mobile devices as input for a spatially structured epidemic model. In fact, mobile devices data can be used to evaluate, in near real-time, the effects of interventions and self-initiated behavioral changes on the mobility of people and to inform large scale epidemic models [4–8]. Here, we use anonymised data provided by a major mobile phone operator in South America (Telefónica Movistar), with a market share of 24.61% as of March 2020.

To characterize the changes of mobility and physical contacts during the outbreak, we used anonymised data from 1.4 million mobile devices (about 22% of the total population in the *comunas* under consideration). We find consistent downward trends coinciding with the NPIs issued by local and national authorities. We estimate that the first set of NPIs issued on 16/03 led to a reduction of about 48% in the number of travels between *comunas*. An additional 17% reduction is observed with the introduction of the full lockdown on 15/05. Furthermore, we find that changes in mobility patterns strongly correlate with the economic and development indicators of the *comunas*.

We develop a stochastic mechanistic epidemic model integrating the real-time mobility, physical contacts, and census data. The model suggests that the full lockdown, while causing a modest additional decrease in mobility and physical contacts with respect to the NPIs already in place, was decisive in bringing the epidemic under control. This relatively small additional decrease in mobility and contacts was enough to push the effective reproductive number below the critical value of 1, a clear example of the threshold effects characterizing epidemic dynamics on structured mobility networks [9]. We estimate that the full lockdown prevented an additional 34.7% (95% CI: [27.2%, 44.1%]) increase in the total number of deaths. Additionally, we estimate the critical impact of the timing of the full lockdown through counterfactual scenarios: an additional week of delay would have corresponded to an 18.1% (95% CI: [6.0%, 34.0%]) more intense incidence peak according to our estimates. The model captures the heterogeneous burden of COVID-19 across *comunas*, observed in the epidemiological data reported by the national surveillance. This highlights how communities exhibiting systemic social disparities are affected in a differential way by government-mandated NPIs due to challenges faced in reducing their mobility and contacts, raising the issue of health disparities in the management of emerging infectious diseases such as COVID-19.

## 2 Results

We evaluate the effects of NPIs policies, government-mandated mobility limitations by integrating mobile phone data and an epidemic model. We identify three phases of the epidemic management in Santiago: i) before 16/03 (business as usual, baseline) ii) between 16/03 and 15/05 (first set of NPIs interventions), and iii) after 15/05 (full lockdown). For convenience, we will refer to the period 16/03-15/05 as the *partial lockdown* and to the period after 15/05 as the *full lockdown*. It is important to notice how the timeline of interventions is fairly complex. It includes night curfews, dynamic quarantine, and lockdowns restricted to a few *comunas* across the region studied here and in other parts of Chile [2]. However, as we see below, the data suggest that those measures did not have a significant impact on people’s behaviors, thus for simplicity we consider the two main sets of NPIs only. We characterize the three phases outlined above in terms of a) mobility among *comunas*, and b) contacts reduction between individuals. Commuting describes the (varying) rates at which people travel among different *comunas*, while contacts reduction parameters estimate to what extent physical contacts drop in each *comuna* (more details in the Materials and Methods section).

### 2.1 Effects of NPIs and Social Inequalities

In Fig. 1A we provide an overview of mobility in Santiago during the period of study. As a proxy for general mobility, we consider the number of devices visiting a *comuna* that is different from their home one (see Material and Methods section). We observe a sharp drop following the first set of interventions on 16/03. Afterwards, mobility remains fairly constant until the introduction of the full lockdown on 15/05, when we observe an additional ∼15% decrease. As we will show below, this intervention represented an important tipping point of the epidemic in Santiago.

**Figure 1:**
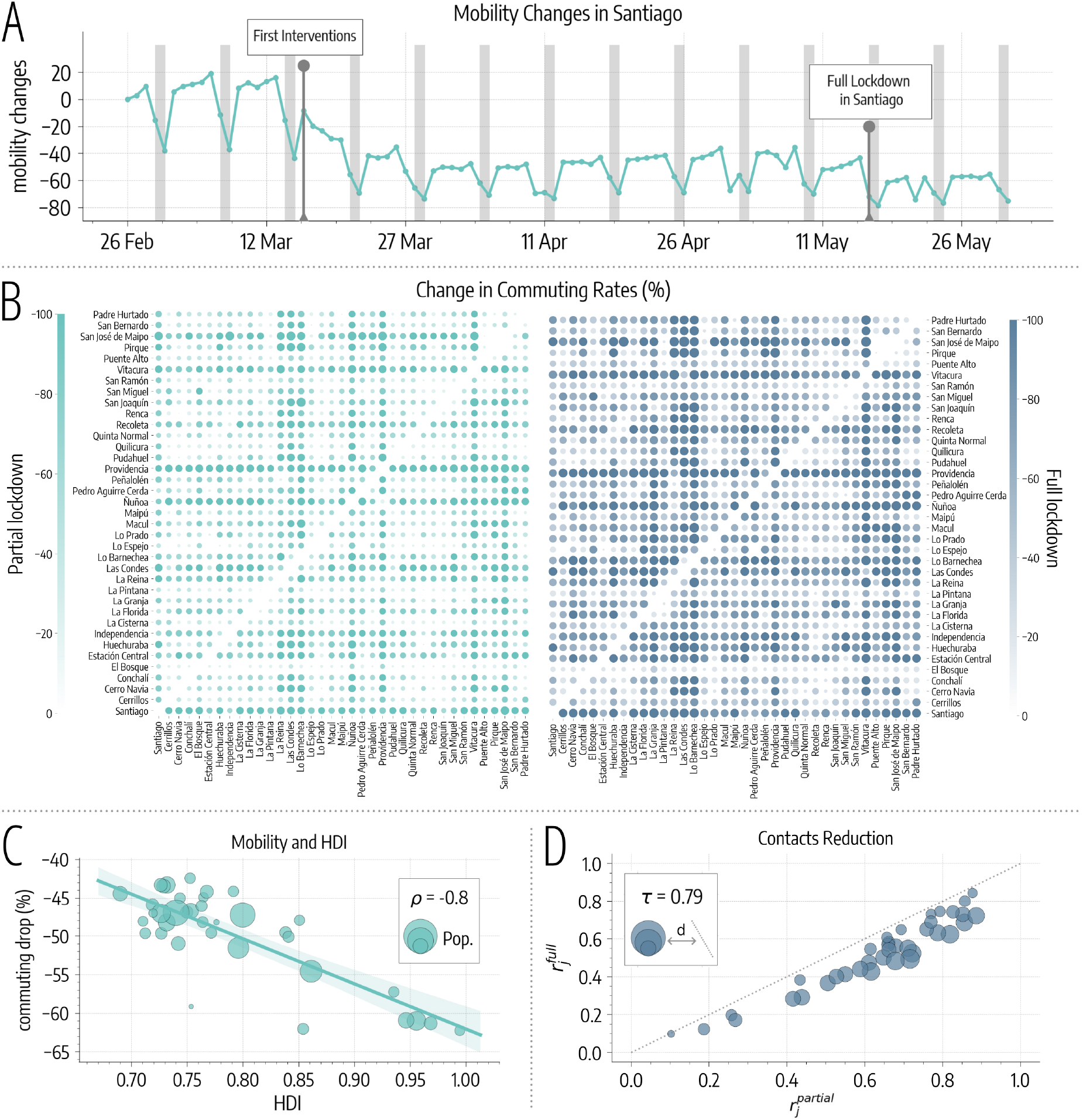
Mobility and contacts changes in Santiago. A) Overview of mobility changes, we consider the number of devices visiting a *comuna* different from their home one as a proxy for general mobility (grey areas represent weekends). Changes are expressed as percentages with respect to 26/02. B) Percentage changes in commuting rates (with respect to commuting before 16/03). On the left drop in commuting after the partial lockdown, on the right after the full lockdown. Color and dots size are scaled according to the magnitude of the change. C) Average percentage commuting decreases after 16/03 versus HDI of different *comunas*. D) Scatter plot of contacts reduction parameters during partial 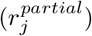 and full 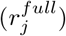 lockdown. Dots size is proportional to the distance from the diagonal (bigger dots indicate *comunas* where contacts decreased more after the full lockdown).

More in detail, we represent changes in commuting among *comunas* in Fig. 1B. The partial lockdown causes an average drop of about 48%, while, with the introduction of the full lockdown, commuting drops by 65% with respect to the baseline. For each *comuna* we also consider the mean percentage decrease in commuting after 16/03 and compare it with the Human Development Index (HDI), a coefficient that measures key aspects of human development, such as life expectancy, education, and per capita income [10]. In Fig. 1C, we observe that a greater decrease in mobility is generally associated with a higher HDI (Pearson correlation coefficient *ρ* = −0.80, *p* < 0.001). The same trend is observed in the absolute change of mobility (see the Supplementary Information) suggesting that wealthier and more developed *comunas* became more isolated after the interventions. This result is in line with previous studies that showed how changes in mobility patterns following government-issued interventions, and the extent to which people can afford social distancing, vary across different socio-demographic groups [11].

In Fig. 1D, we represent contacts reduction parameters. Across the board, contacts drop by 36% with the first set of NPIs policies and by an additional 11% with the lockdown. Since all points are below the diagonal, we conclude that, with the introduction of the full lockdown, contacts decrease further in all *comunas*. Also, the decrease is consistent with the existing reductions after the partial lockdown. Indeed 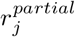 and 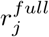 show a high significant correlation (Kendall rank correlation coefficient *τ* = 0.79, *p* < 0.001).

### 2.2 The Spread of COVID-19 in Santiago

We use the mobility data to develop and to inform a stochastic mechanistic metapopulation epidemic model (see Materials and Methods for details) and simulate the spread of COVID-19 in the *comunas* of Santiago. The model is calibrated on official surveillance data and takes as initial seeding the realistic projections of active cases on March 1^st^, 2020 in the Metropolitan area of Santiago from Ref. [7].

We use an Approximate Bayesian Computation (ABC) approach [12, 13] (see details in the Materials and Methods section) to find the posterior distribution of the reproductive number in Santiago (median *R*_0_=2.66, 95% CI: [2.58, 2.72]), which is in line with previous findings that identify the value of *R*_0_ of SARS-CoV-2 to be in the range between 2 and 3 in different countries [14–16]. In Fig. 2A we report the number of weekly deaths projected by the model together with official figures (used for calibration). The two time series show a good agreement with a high correlation (*ρ* = 0.99, *p* < 0.001) and a median absolute percentage error of 12%. Interestingly, the agreement between data and model starts to deviate in the last two data points (end of July early August). We can speculate that, at least in part, this might be due to a lockdown fatigue. In fact, while the official restrictions were relaxed later (mid of August), a decrease in compliance, linked to the reduction of cases/deaths, could have taken place earlier. The period is outside our current data coverage. Hence, we leave testing such hypothesis to future work.

**Figure 2:**
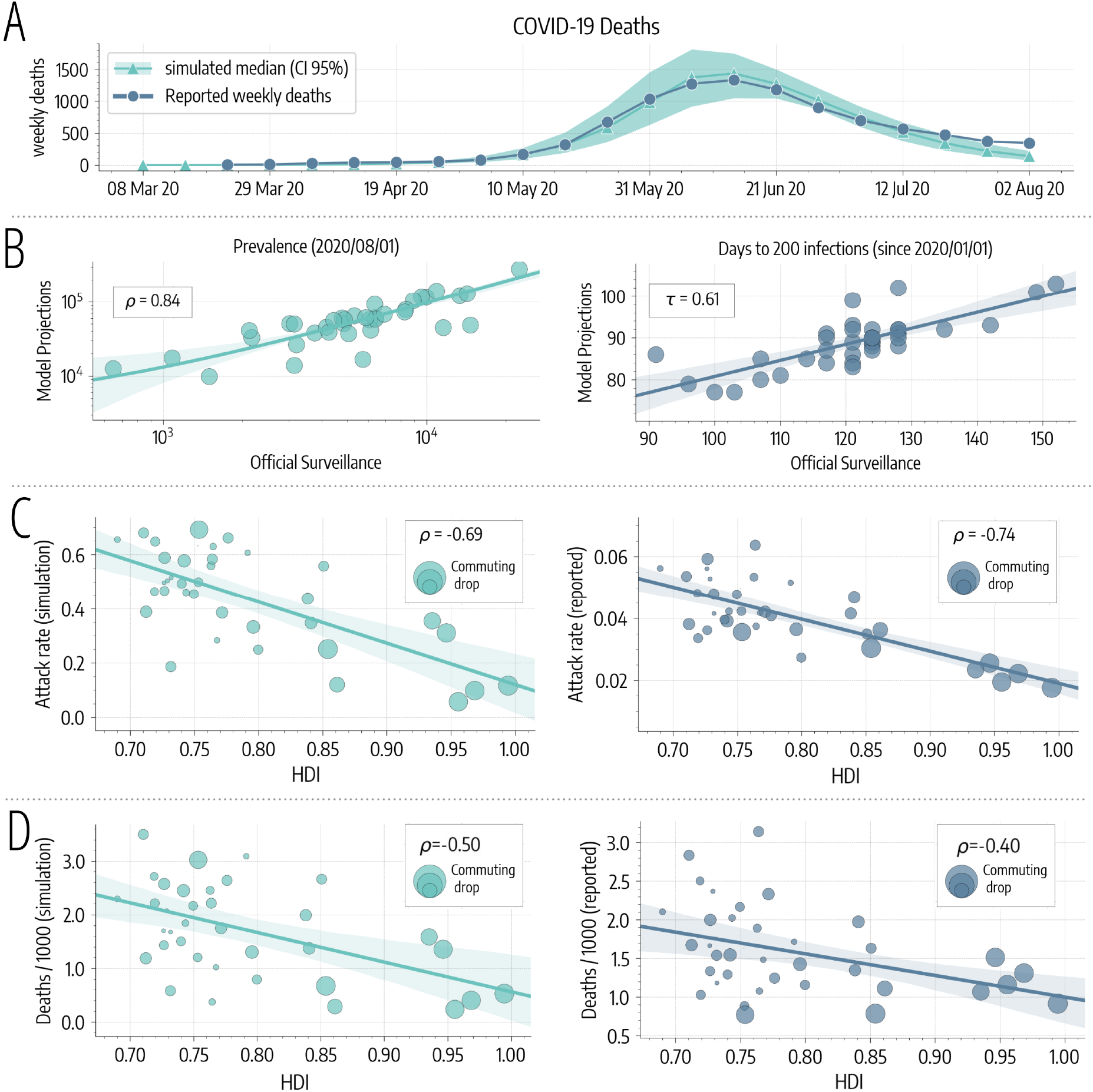
COVID-19 spreading in Santiago. A) We represent the simulated and reported weekly deaths used for model calibration. B) Left, scatter plot of reported versus simulated cases as of 2020/08/01. Right, scatter plot of days (since 2020/01/01) needed to reach 200 in each *comuna* as reported by official surveillance and as projected by our model. C) Scatter plot of HDI versus attack rate as of 2020/08/01 in different *comunas* as projected by our model (left) and as reported by official surveillance (right). Size of dots are scaled according to commuting drops after 16/03 (bigger bullets indicate bigger decreases in commuting). D) Scatter plot of HDI versus deaths per 1, 000 as of 2020/08/01 in different *comunas* as projected by our model (left) and as reported by official surveillance (right). Size of dots are scaled according to commuting drops after 16/03.

As of August 1^st^, 2020, the median projected fraction of infected individuals in the area under study is 38.7% (95% CI: [35.1%, 41.6%]). This estimate is about tenfold the official reported figures. We are not aware of publicly available seroprevalence studies that we can use as a comparison and validation. For this reason, we can only consider qualitative evidence hinting that the Chilean outbreak has affected a significant fraction of the population. For example, the share of positive COVID-19 tests peaked at 59.10% on June 18^th^, and in the Santiago Metropolitan Region the occupation of ICU beds reached almost saturation level (95%) in May. Also, a recent epidemiological study aimed at characterizing the first wave in Chile showed significant under-reporting of symptomatic cases (around 50%) based on estimates of the Case Fatality Rate [2]. Finally, previous seroprevalence studies conducted, for example, in the United States [17] and Spain [18], showed that the actual number of COVID-19 infections is several times (factors vary from 4 to 20) those reported by the official surveillance. As a sensitivity check, we repeated the calibration considering the upper limit of the 95% credible interval for the infection fatality rate from Ref. [19]. This leads to a projected median prevalence of 28.3% (95% CI: [23.5%, 32.3%]) but to a sensibly worse fit of the data (*ρ* = 0.82, median percentage error of 49%).

Projected cases present a good significant correlation with official numbers as can be observed in Fig. 2B (*ρ* = 0.84, *p* < 0.001). Besides, we compare the dates when 200 infections have been reached in different *comunas* according to our model and official surveillance, finding a significant correlation (Kendall rank correlation coefficient *τ* = 0.61, *p* < 0.001). Similar results are found considering instead the dates when 50, 100, and 500 cases have been reached (see the Supplementary Information). Interestingly, the same dates estimated through modeling are much earlier, hinting that many of the infections in the initial phase of the spreading went unreported. In Fig. 2C we show the attack rates versus the HDI of different *comunas*. We find a strong correlation between attack rates computed on officially reported cases and HDI (*ρ* = −0.74, *p* < 0.001), providing evidence that wealthier *comunas* experienced significantly smaller outbreaks. In addition, the very same picture emerges from our modelling results. Indeed, simulated attack rates present a high significant correlation with HDI (*ρ* = −0.69, *p* < 0.001). Finally, in Fig. 2D we show the number of deaths per 1, 000 versus the HDI of different *comunas*. We find a significant correlation between HDI and both simulated (*ρ* = −0.50, *p* < 0.002) and officially reported (*ρ* = −0.40, *p* < 0.02) deaths, hinting that wealthier *comunas* experienced also a smaller burden in terms of casualties. We note that the correlations obtained in this case are lower than those found previously for the attack rates. This may be due to the interplay between diverse age distributions and age-dependent mortality rates. Indeed *comunas* with higher HDI have a higher mean age and the infection fatality rate for COVID-19 is significantly higher in older age brackets.

### 2.3 Counterfactual Scenarios

To assess the impact of heterogeneous responses to the spreading we run a hypothetical scenario in which commuting and contacts decrease uniformly across *comunas*, starting on 16/03. More in detail, we apply to all *comunas* the average reduction in commuting and contacts observed for the 4^th^ quartile of HDI (i.e. 25% *comunas* with higher HDI). See the Supplementary Information for more details. According to our simulations, this leads to a significant decrease of cases and deaths: −83.8% (95% CI: [−77.6%, −88.6%]) fewer cases and −70.5% (95% CI: [−55.0%, −80.9%]) fewer deaths as of May 15^th^, the date when the full lockdown was enforced. Interestingly, the uniform reduction we are imposing implies a relatively modest additional decrease to commuting and contacts with respect to the ones estimated through mobile phone data. In this hypothetical scenario, with the partial lockdown commuting rates drop by 55% and contacts by 49% (versus respectively the 48% and the 36% estimated in our main analysis). Although such homogeneous reduction across *comunas* is a theoretical exercise that does not consider complex socio-economic constraints, that go from the collective need to keep key supply chains active to the individual imperative to feed their own family, it crystallizes the dramatic effects of inequality on disease spreading on the one side, and it shows the positive benefits of equal, early, and strong responses on the other.

We also use the model to investigate counterfactual scenarios aimed at estimating the impact of NPIs on the spread of COVID-19 in Santiago. As a first counterfactual scenario, we simulated the epidemic in the absence of a full lockdown. From Fig. 3A we observe that this leads on average to a 21.6% (95% CI: [7.5%, 41.3%]) more intense incidence peak and 34.7% (95% CI: [27.2%, 44.1%]) more deaths. To estimate the impact of the timing of the full lockdown, we run simulations where we anticipate or delay it up to four weeks. According to results in Fig. 3B, an earlier lockdown implies a less intense incidence peak (from around −20% to −35%). It is interesting to note, however, that a delay of 1-2 weeks has a very similar effect of no intervention at all in terms of incidence peak intensity. More specifically, one week delay causes a 18.1% (95% CI: [6.0%, 34.0%]) while two weeks delay cause a 21.6% (95% CI: [7.4%, 41.1%]) more intense incidence peak. The timing of the full lockdown also has a significant effect on the number of deaths. According to our estimates in Fig. 3B, just one week of delay implies a 7.7% (95% CI: [1.3%, 13.7%]) increase in mortality.

**Figure 3:**
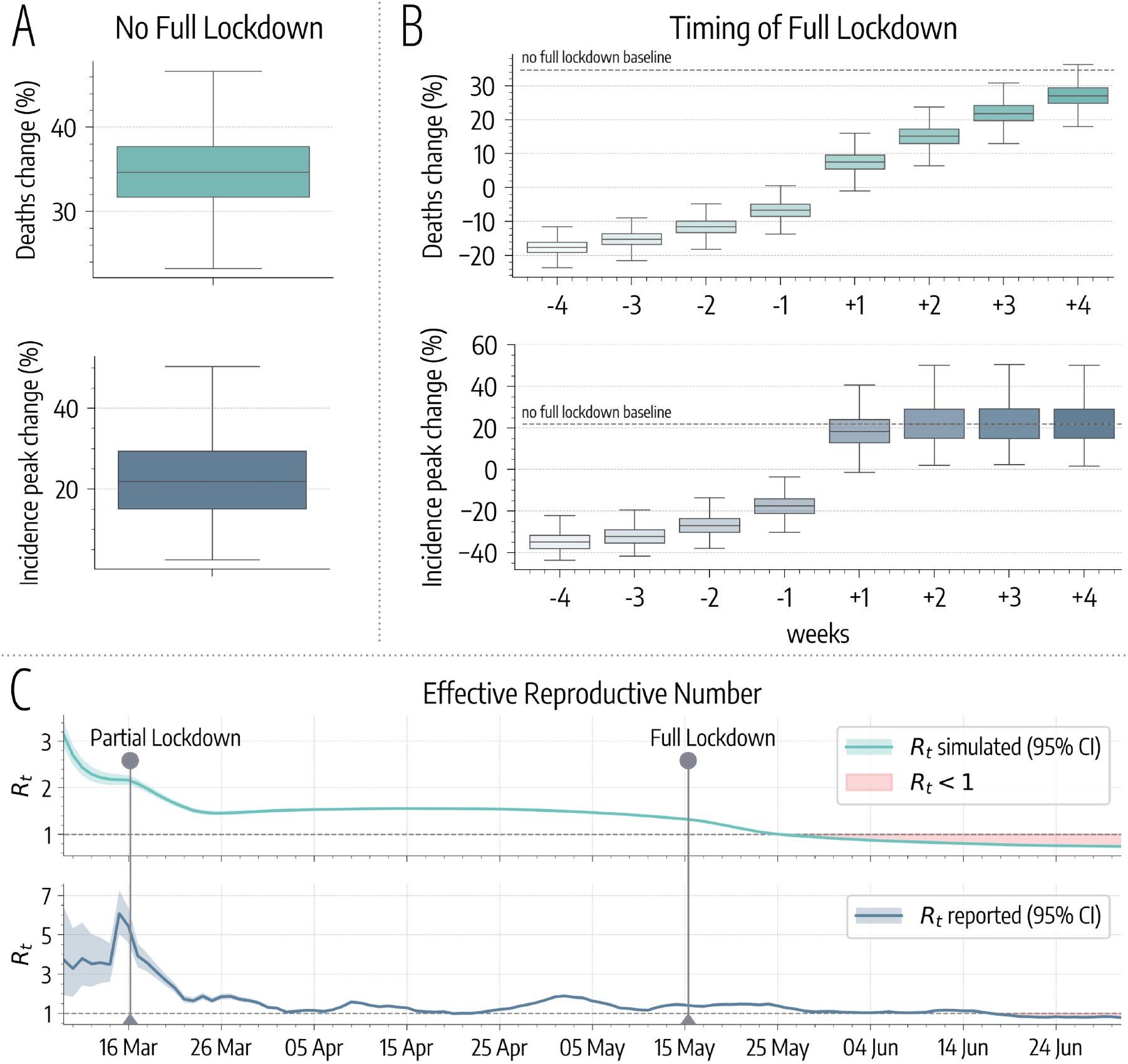
Impact of non-pharmaceutical interventions on COVID-19 spreading. A) Model estimates of percentage increases in deaths and incidence peak intensity without the implementation of the full lockdown. B) Model estimates of percentage changes in deaths and in incidence peak intensity moving the date of the full lockdown of −4/ + 4 weeks. C) Effective Reproductive Number *R*_*t*_ estimated on simulated and officially reported cases. The two time series show a high positive Pearson correlation coefficient (*ρ* = 0.78, *p* < 0.001).

### 2.4 Effective Reproduction Number

In Fig. 3C we show the evolution of the effective reproduction number *R*_*t*_ estimated using the method from Ref. [20] on the simulated and the official reported incidence. In the simulated *R*_*t*_ time series we observe the two discontinuities after the implementation of government-issued NPIs. However, we note that the partial lockdown had the sole effect of slowing down the epidemic. Indeed, after March 16^th^ the estimated *R*_*t*_ is still greater than 1. After the full lockdown, instead, *R*_*t*_ was pushed below 1 making the containment possible. This is visible both in the simulated and the reported time series. This result underlines the importance of the full lockdown that, despite causing a relatively small effect on mobility, had a decisive role in bringing the outbreak under control. It is worth stressing this result. The full lockdown constituted a key tipping point for the evolution of the epidemic pushing the reproductive number below its critical threshold. A similar finding has been recently reported for the evolution of the pandemic in Germany [21]. Indeed, also in that context, only the subsequent compounding of interventions was able to bring the reproductive number below one thus curbing the spreading the virus.

## 3 Discussion

The analysis presented here shows that the effects of NPIs issued by the government strongly correlate with a measure of human development, such as the HDI. In particular, *comunas* with higher HDI were able to reduce more significantly their mobility. This, in turn, is reflected in both data and modeling estimates by a lower burden of COVID-19 (cases, deaths) in the *comunas* characterized by a higher HDI. The combination of these results raises policy-making concerns. Indeed, while lockdowns are unquestionably effective in mitigating the epidemic activity, they may as well augment social and health inequalities, penalizing more vulnerable communities. Other studies have found that mobility restrictions unequally affected different regions of France [8], Italy [22], United States [11], Colombia, Mexico, and Indonesia [23] with a higher income being associated to a larger capacity to afford social distancing. Furthermore, observations in the United States [11, 24–26], Singapore [27] and the UK [28, 29] show that socio-economic inequalities are linked to worst health outcomes during the current pandemic.

Our data-driven analysis also shows that the timeliness of NPIs is just one variable influencing the outcome of the mitigation effort. The case of Santiago is emblematic. NPIs were introduced early respect to other countries. Only two days after the first 50 confirmed cases. For comparison, Denmark introduced measures after five, Austria after nine, Italy and Germany after fifteen days of reaching that threshold [30]. These measures were followed by a considerable reduction in mobility and contacts, but cases soared anyway in the metropolitan area. According to our analysis, the first set of NPIs significantly slowed down transmissions but not enough to stop the epidemic. It was only after the introduction of the second additional lockdown that the Santiago outbreak was brought under control. This indicates that earlier implementation of more stringent NPIs may be beneficial in quickly mitigating the outbreak without extending for a long time policies that potentially might unequally affect communities.

The present work comes with limitations. First, we focused only on the epidemic evolution within the Santiago Metropolitan Area and we overlooked both national and international importations after March 1^st^, 2020. While it is reasonable to assume that after this date the epidemic was largely sustained by the internal spreading (especially considering the various restrictions on international and national mobility), we acknowledge this as a possible limitation. Second, compared to other approaches [31–33], we considered a relatively simple disease dynamics. Lastly, we acknowledge that our mobile phone users’ sample was not selected to be representative of the whole population, but we know that Telefónica Movistar data can well represent the different socio-demographic groups of Santiago [34].

Overall, our study characterizes the unfolding of SARS-CoV-2 in one of the largest metropolitan areas in South America; a region that so far has received far less attention than others. It quantifies the unequal effects across communities of behavioral changes introduced by governmental measures as well as individual (re)actions and provides evidence that even small delays in the implementation of NPIs can have a significant impact on the unfolding of the epidemic.

## 4 Materials and Methods

### 4.1 Measuring Mobility and Contacts

In this work, we use phone data in the form of eXtended Detail Records (XDR). This stream records every interaction (e.g. packet request) between devices and antennas. An entry in our dataset can be formalized as a tuple ⟨*d, t, a*⟩ indicating a packet request to antenna *a* made by device *d* at time *t*. We approximate the position of *d* with that of the antenna, which has a fixed latitude and longitude. We also assign a home antenna to each device by finding the most active antenna during night hours. Finally, we assign antennas to correspondent *comunas* according to their position. The dataset includes data for the period 2020/02/26 − 2020/06/01 for 1.4 million devices which correspond to the 22% of the total population in the area considered. To preserve the privacy of device owners, we analyze and display only anonymous and aggregated results. Furthermore, no other information about the users (i.e. gender, age etc..) was used nor available.

As specified in the previous sections, we characterize the three phases of the outbreak in terms of commuting and contacts reduction. Commuting is measured considering the fraction of devices traveling between *comunas*. Formally, for each day *t* we build a commuting rate matrix Σ(*t*) ∈ ℝ^*N* ×*N*^ whose element *σ*_*ij*_(*t*) is the fraction of devices living in *comuna i* that visited *j* on day *t*. We average these daily rates during the three phases to obtain three distinct matrices describing commuting i) before any restrictions, ii) during the partial lockdown, and iii) during the full lockdown.

It is important to notice that the data does not provide direct information about physical contacts between users. Having the privacy of the users in mind, and considering the various non-trivial assumptions one would need to make, we do not attempt to estimate/infer such contacts. Instead, we focus on a metric that allows us to capture the variation before and after the various interventions. As we describe below, the epidemic model considers a homogeneous mixing approximation within each subpopulation (i.e. comuna) hence the only important variable is an estimate of contacts reduction rather than the actual contacts. To this end, contacts reduction is estimated by looking at the variation in the number of users co-located in the same antenna. Each antenna *a* in comuna *j* has a resident population 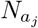. On day *t*, the total number of visitors from the same *comuna* is 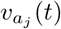. Assuming homogeneous mixing, the maximum number of contacts in antenna *a* is 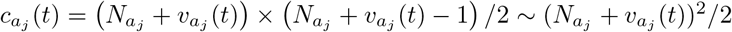. Then, we assume the reduction of contact during the partial and full lockdown to be equal to:

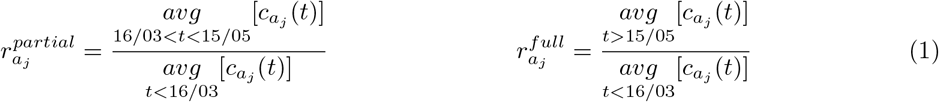

In other words, the reduction of contacts during the partial and full lockdown is considered as the variation of the maximum number of contacts before and after each intervention. Finally, we aggregate at the level of *comunas* taking the median of these quantities over all antennas located in the same *comuna*.

### 4.2 Modeling the spread of COVID-19 in Santiago

The model used in this work to simulate the spread of COVID-19 is largely inspired by the Global Epidemic and Mobility Model (GLEAM) [35, 36]. In this section, we present the conceptual framework but a full mathematical description is provided in the Supplementary Information.

The *comunas* of Santiago are represented as single subpopulations in a metapopulation network. Inside each one, we divide individuals into *K* = 16 five-year age brackets respecting the demographic of different *comunas* [37] and we use the country-specific contact matrix from Ref. [38] to define the rates at which different age groups mix with each other. Individuals are also divided into compartments according to their health status. We consider a SLIR (Susceptible, Latent, Infectious, Removed) compartmentalization setup. A similar approach has been used in several modeling studies in the context of COVID-19 [7, 39, 40]. Interacting with Infectious, Susceptibles move to the Latent stage in which they are not infectious yet. Only after the incubation period, Latent become Infectious. Lastly, Infectious transit to the Removed compartment at a rate inversely proportional to the infectious period.

Individuals can get the infection interacting with infected in their home and in other connected metapopulations. To model this aspect, we consider the commuting network previously introduced to describe the *coupling* (i.e. the strength of connection) between *comunas*. Technically, we define a “force of infection” 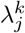 that expresses the infection rate for individuals in age group *k* residing in *comuna j*:

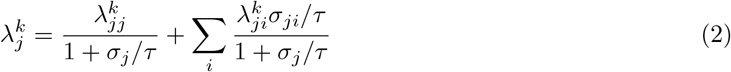

Where *τ* set the time scale of commuting (here *τ* ^−1^ = 1/3 *day*) and *σ*_*j*_ = Σ_*i*_*σ*_*ji*_ is the total commuting rate of population *j*. In particular, the first term in Eq. 2 represents the contribute from active infections in *comuna j*, and the summation from cases in other connected *comunas i* (see Supplementary Information for full details).

Government interventions are implemented by changing the commuting network on 16/03 (partial lockdown commuting) and again on 15/05 (full lockdown commuting) to reflect the corresponding variations in mobility. Similarly, on these dates, we multiply the age contact matrix of each *comuna* by 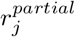 and 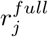, respectively. We do not explicitly account for school closure since its effect is already included into the changes inferred from mobile devices data.

The model is fully stochastic and transitions among compartments are simulated through chain binomial processes. In the main text we present results for an incubation period of 4 days and an infectious period of 2.5 days, which imply a generation time *T*_*G*_ = 6.5 days, in line with current estimates [41, 42]. We simulate deaths considering the estimates of the Infection Fatality Rate from Ref. [19] and a delay Δ after the transition to the Removed compartment.

Initial seeding is done using the projections of active cases on March 1^*st*^, 2020 in the Metropolitan area of Santiago from Ref. [7] and assigning infections to different *comunas* proportionally to the population distribution. The calibration is performed on weekly deaths using an Approximate Bayesian Computation (ABC) Rejection method [12, 13]. At each step of the ABC algorithm, a set of parameters *θ* is sampled from a prior distribution and an instance of the model is generated using these parameters. Then, an output quantity *E*′ of the model is compared to the corresponding real quantity *E* using a distance measure *s*(*E*′, *E*). If this distance is greater (smaller) than a predefined tolerance *ϵ*, then the sampled set of parameters is discarded (retained). After a sufficient number of iterations, the distribution of accepted sets will approximate the posterior distribution of parameters *P* (*θ, E*) given the evidence *E* from the data. In this work, we set a flat uniform prior on the parameters (*R*_0_ ∈ [2, 4] in steps of 0.02 and Δ ∈ [14, 21] days in steps of 1 day) and we perform calibration using the median absolute percentage error as a distance metric with a tolerance of 20% on weekly deaths. We run 140, 000 iterations which correspond to about 200 stochastic realizations for each possible parameters set. We use the official data issued by the Department of Statistics of the Chilean Minister of Health [43]. We consider both COVID-19 “confirmed” and “suspected” deaths to perform the calibration, in the Supplementary Information we support this decision showing that considering only confirmed COVID-19 deaths we still obtain a significant anomaly in mortality. Model projections are produced sampling parameter sets directly from the posterior distribution and generating an ensemble of trajectories. In this work we generate model estimates sampling 5, 000 sets on which we compute median and confidence intervals.

## Data Availability

The raw data analysed in the study are not publicly available due to privacy reasons. All the aggregated mobility data needed to run the model are available. Analysis of the anonymised mobile phone data was performed on mobile operator's systems without transferring it outside. Only aggregated mobility patterns across municipalities were provided to researchers outside Chile and only these have been used for the analyses presented here

https://github.com/ngozzi/covid19-santiago

## Acknowledgements

All authors thank the High Performance Computing facilities at Greenwich University. M.T. acknowledge support from the Lagrange Project of the Institute for Scientific Interchange Foundation (ISI Foundation) funded by Fondazione Cassa di Risparmio di Torino (Fondazione CRT). M.T. acknowledges support from EPIPOSE - “Epidemic intelligence to minimize COVID-19’s public health, societal and economical impact” H2020-SC1-PHE-CORONAVIRUS-2020 call. L.F. thanks Víctor Navarro, and acknowledges the funding and support of Telefónica R&D Chile and CISCO Chile. N.G. acknowledges support from the Doctoral Training Alliance. M.C. and A.V. acknowledge support from Google Cloud and Google Cloud Research Credits program.

## Authors Contribution

N.G., M.T., L.F., N.P. and A.V. designed the study. N.G. and N.P. analyzed the data. N.G. implemented and run the epidemic model. L.F. mined and provided the anonymised mobile phone data. M.C. provided the importation data to initialise the model. All authors interpreted the results, wrote and approved the manuscript.

## Funding Acknowledgements

A.V. reports grants and personal fees from Metabiota inc., outside the submitted work; M.C. report grants from Metabiota inc., outside the submitted work; M.T. report personal fees from GSK, outside the submitted work. No other relationships or activities that could appear to have influenced the submitted work.

## Data Availability

The raw data analysed in the study are not publicly available due to privacy reasons. All the aggregated mobility data needed to run the model are available at https://github.com/ngozzi/covid19-santiago. Analysis of the anonymised mobile phone data was performed on mobile operator’s systems without transferring it outside. Only aggregated mobility patterns across municipalities were provided to researchers outside Chile and only these have been used for the analyses presented here. The study was deemed exempt (IRB #20-10-05) by the Northeastern University Internal Review Board.

## A Supplementary Information

### A.1 Sensitivity Analysis

In this section we provide a sensitivity analysis on the data used for model calibration, on model parameters, on the correlation between mobility changes and sociodemographics, and finally on the number of days needed to reach *N* infections according to our estimates and official surveillance in different *comunas*.

#### Deaths Data for Calibration

We calibrate the model using COVID-19 deaths data issued by the Department of Statistics of the Chilean Minister of Health (DEIS) [43]. The dataset includes both “confirmed” (i.e. supported by a clinical test, ICD code: U07.1) and “suspected” deaths (i.e. supported only by symptoms, ICD code: U07.2). In Fig. 4A we compare 2020 all-cause deaths with historical median and 95% confidence intervals. We observe that, excluding only COVID-19 confirmed from 2020 deaths we still obtain a significant anomaly. Excluding also suspected, instead, we obtain a trend that is within confidence intervals. For this reason, we decide to calibrate the model considering both COVID-19 confirmed and suspected deaths.

**Figure 4:**
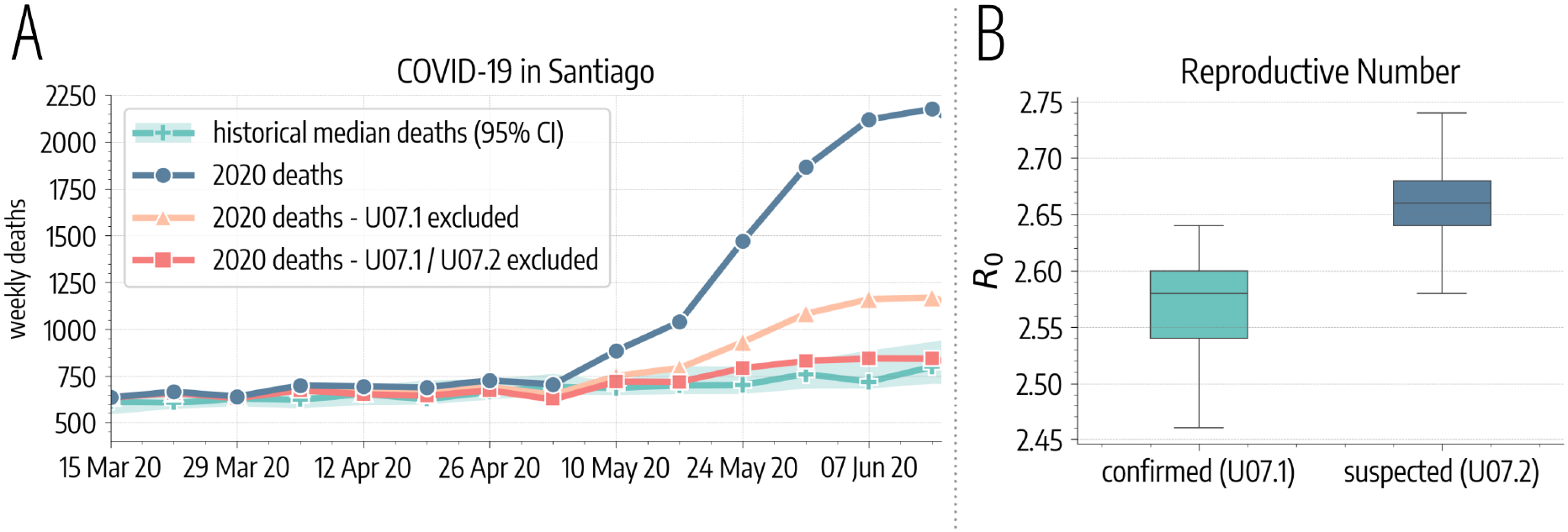
Impact of COVID-19 on mortality rate in Santiago. A) We plot 2020 all-cause weekly deaths in the *comunas* of Santiago and the historical median with 95% confidence interval computed on 2010-2019 data. We report also 2020 deaths time series excluding only COVID-19 confirmed (ICD code: U07.1) and also suspected (ICD code: U07.2) deaths. B) *R*_0_ estimates considering only confirmed and also suspected COVID-19 deaths.

Here, we perform also calibration considering only confirmed COVID-19 deaths. In Fig. 4B we observe that the estimates for *R*_0_ obtained in this case do not change significantly (median *R*_0_ of 2.58, 95% CI: [2.50, 2.64]). Moreover, this approach leads to a worse fit of the data (median absolute percentage error of 28% with respect to the 12% obtained considering also suspected deaths).

#### Incubation and Infectious Period

In the main text we considered an incubation period (*T*_*IC*_) of 4 days and an infectious period (*T*_*IF*_) of 2.5 days. Here, we run simulations where we vary these parameters and we observe the impact on our findings. In particular, we consider estimated *R*_0_, prevalence and increase in deaths without the full lockdown for longer incubation (*T*_*IC*_ ∈ [5, 6] days) and infectious periods (*T*_*IF*_ ∈ [3, 3.5] days). In Fig. 5 we represent results of the sensitivity analysis. From Fig. 5A we observe that the median *R*_0_ for the different parameters explored remains in the range [2, 3]. Also, from Fig. 5B we note that the median projected prevalence as of August 1, 2020 in Santiago varies of at most 5.9%. Finally, the results of the counterfactual analysis in which we do not introduce the full lockdown are very robust across different parameters explored. Indeed, the median projected increase in deaths as of August 1, 2020 varies of at most 3.0%.

**Figure 5:**
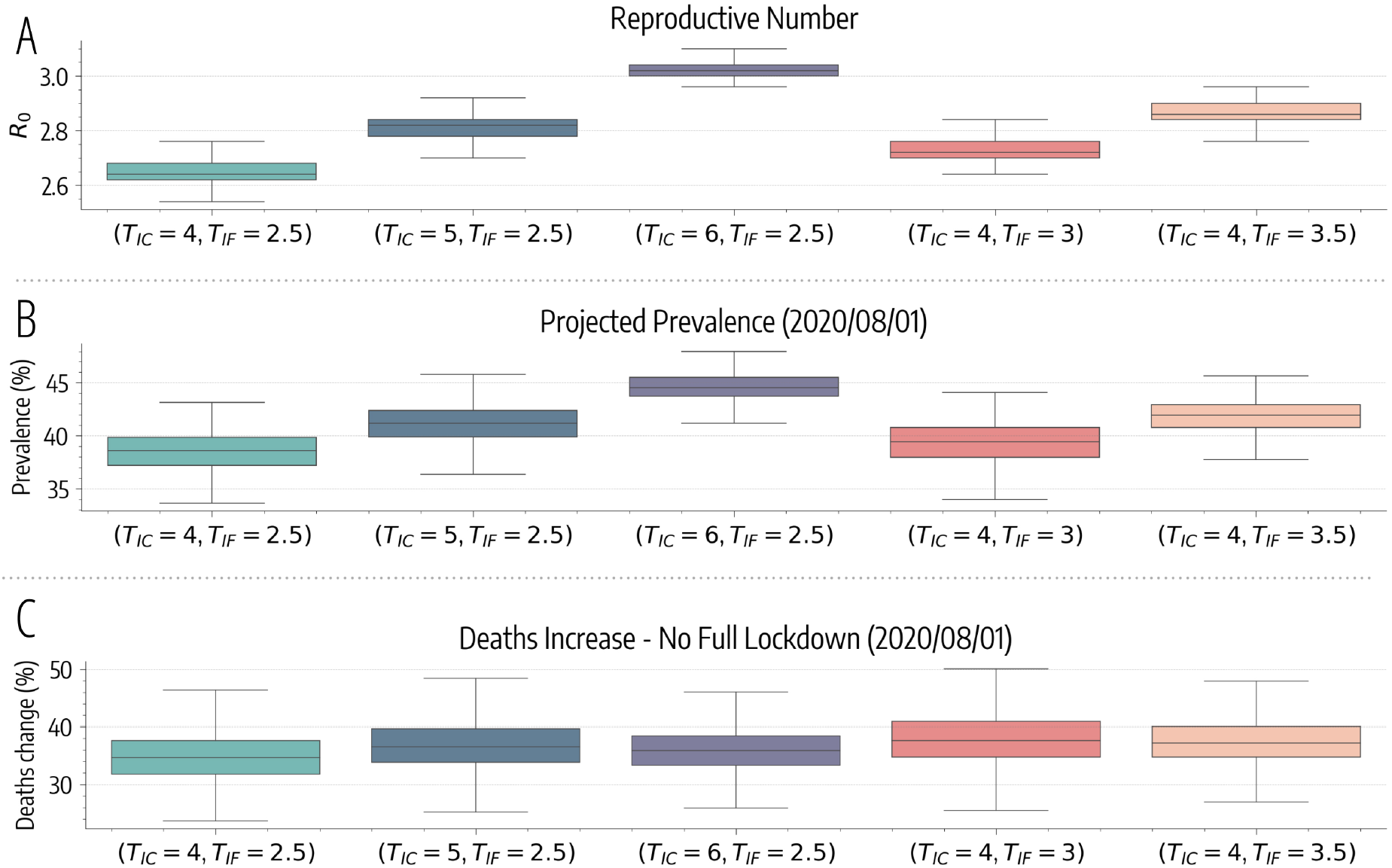
Results of sensitivity analysis. A) Basic reproductive number estimates for different parameters explored in the sensitivity analysis. B) Model projections of prevalence in Santiago as of August 1, 2020 for different parameters. C) Increase in deaths (in percentage) as of August 1, 2020 without the full lockdown for different parameters.

#### Mobility and Sociodemographics

In the main text, we showed that the change in commuting correlates with the Human Development Index (HDI) of different *comunas*. Here, we show that this pattern holds for other economic and development indicators too. In particular, we consider separately the components of the HDI, namely the Life Expectancy Index (LEI), the Education Index (EI), and the Income Index (II) [10]. In Fig. 6 we see that the correlation between these indicators and the average commuting drop after 16/03 of the 37 *comunas* considered is still high, significant, and consistent with the results presented in the main text for the HDI.

**Figure 6:**
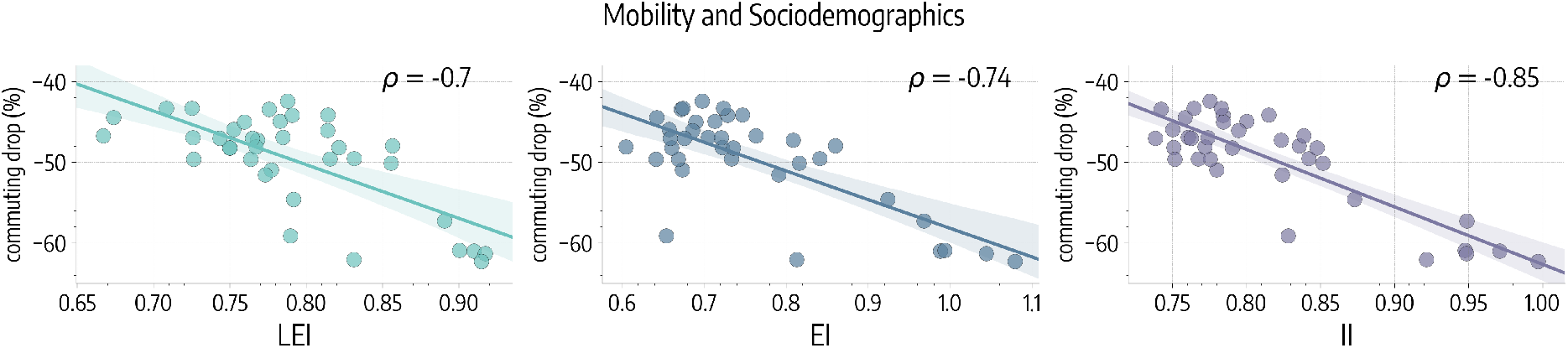
Correlation between mobility and sociodemographics. We represent the scatter plot and the correlations between average percentage drop in commuting after 16/03 and the Life Expectancy Index (LEI), the Education Index (EI), and the Income Index (II).

Also, in the main text we showed the significant correlation between HDI and the average percentage decrease in commuting after March 16^th^, 2020. Here, we show that this finding hold also for absolute commuting decreases. This sensitivity check is needed since a higher percentage decrease does not imply necessarily a lower mobility after the introduction of NPIs (for example, the mobility baseline of wealthier *comunas* can be generally higher). We define the outflow as the average number of outgoing travels per device observed for a *comuna* during a given period. In Fig. 7 we represent (left) the outflow before March 16^th^, 2020 versus the HDI of different *comunas*. We observe no significant correlation (*ρ* = 0.01, *p* = 0.94). On the right, instead we represent the scatter plot of the absolute reduction in outflow before/after March 16^th^, 2020 versus the HDI of *comunas*. In this case we find a strong, positive, and significant correlation (*ρ* = 0.55, *p* < 0.0001), hinting that wealthier *comunas* reduced more their mobility also in absolute terms.

**Figure 7:**
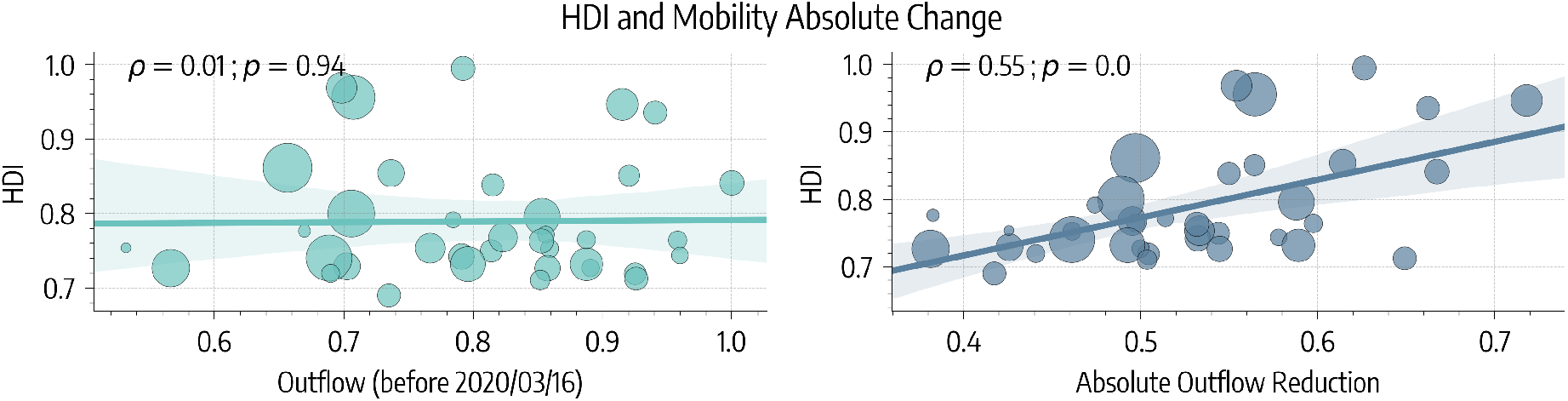
HDI and absolute mobility changes. On the left, scatter plot of outflow (normalized) before March 16^th^, 2020 versus HDI of different *comunas*. On the right, scatter plot of the absolute reduction in outflow after March 16^th^, 2020, versus HDI of different *comunas*. Points size is scaled according to population.

#### Projected cases

In the main text, we assessed the performance of the model comparing via the Kendall rank correlation coefficient the number of days (since 2020/01/01) needed to reach 200 infections in different *comunas* according to our projections and official surveillance. We found a good, significant correlation and we remarked that dates estimated through modeling were generally earlier to those officially reported. In Fig. 8 is shown that this pattern holds also for different numbers of infections. In particular, we consider the amount of days needed to reach 50, 100, 200, and 500 infections.

**Figure 8:**
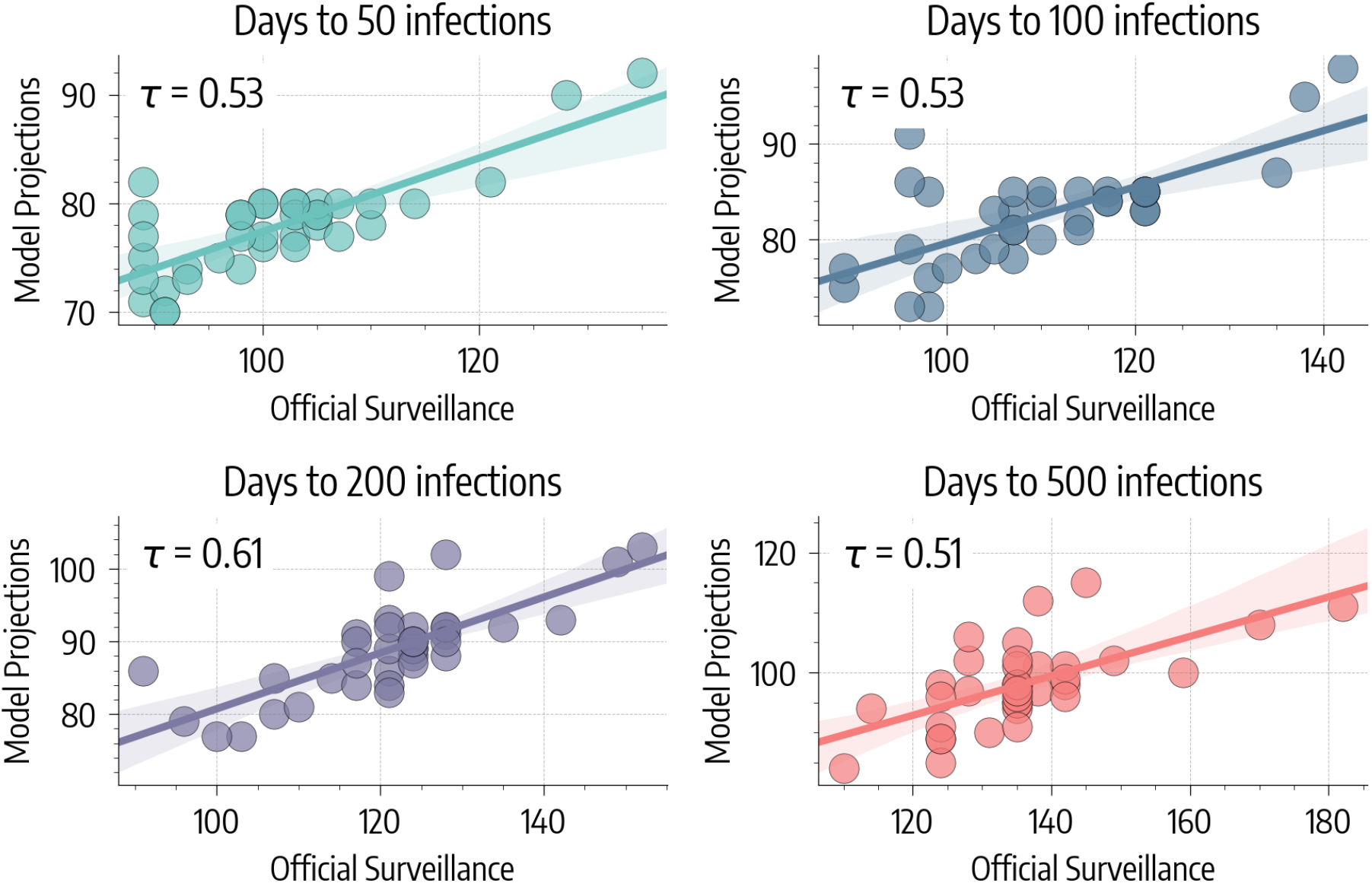
Days to reach N infections in different *comunas*. We consider the number of days since 2020/01/01 needed to reach 50, 100, 200, and 500 infections in different *comunas* as projected by our model and as confirmed by official surveillance.

## B Homogeneous NPIs Effect - Counterfactual Scenario

In the main text we present a hypothetical scenario in which we assign to all *comunas* the average decrease in commuting rates and contacts observed after the partial lockdown for the 4^th^ quartile of HDI (i.e. 25% *comunas* with higher HDI). In Fig. 9A we represent the percentage change in total COVID-19 deaths and cases as of 2020/05/15, date of the 2^nd^ lockdown. We observe that the effect on the spreading is significant: −83.8% (95% CI: [−77.6%, −88.6%]) fewer cases and −70.5% (95% CI: [−55.0%, −80.9%]) fewer deaths. In Fig. 9B we represent the percentage change in COVID-19 cases as of May 15^th^ in different *comunas*. We observe that, in general, the uniform reduction in mobility and contacts brings benefits also to *comunas* with higher HDI. This hints that a more equal distribution of behavioral responses to the NPIs in place would be advantageous not only for the most vulnerable but to the community as a whole.

**Figure 9:**
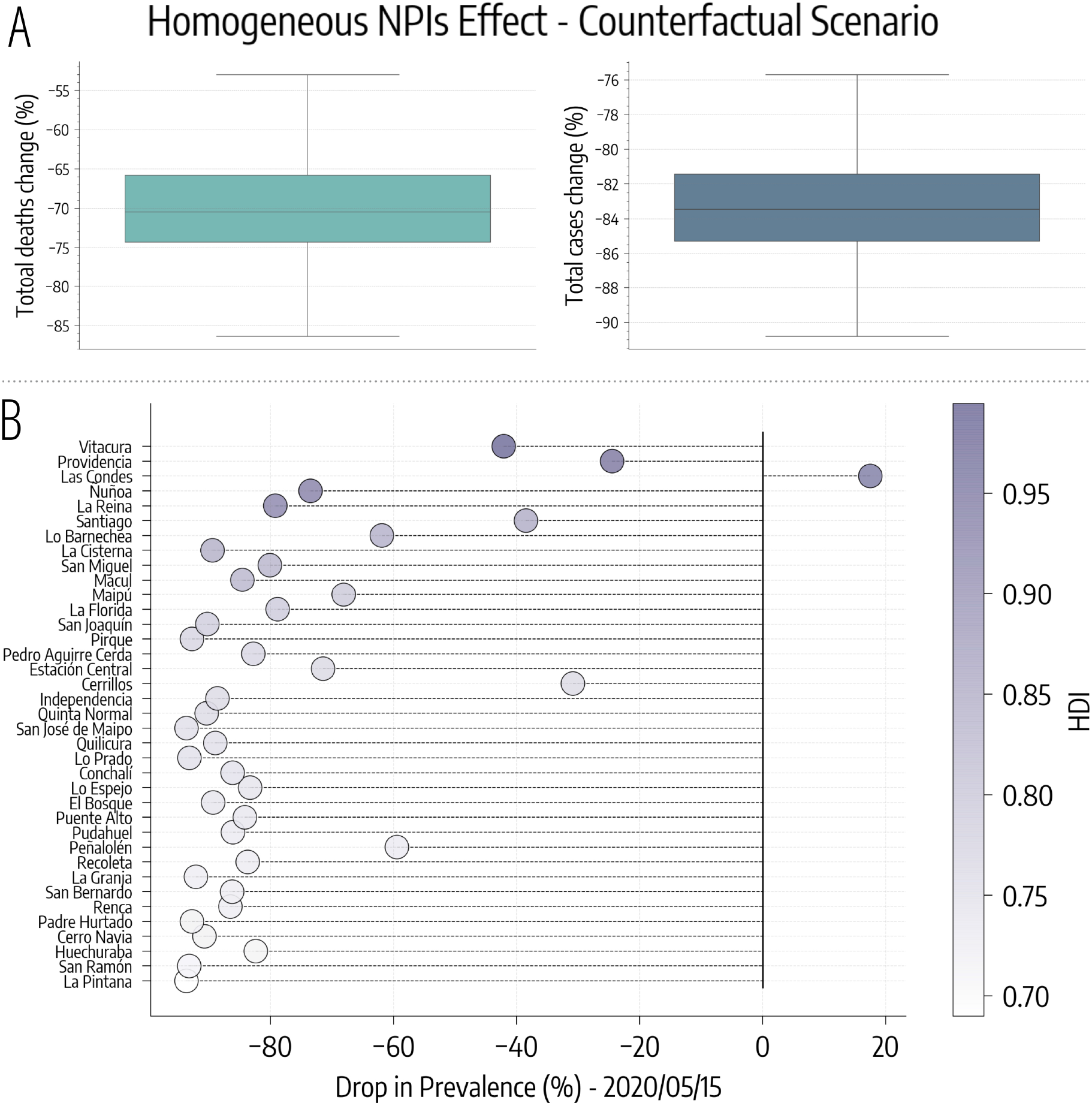
Homogeneous NPIs Effect - Counterfactual Scenario. A) Drop (%) in total number of COVID-19 deaths and cases as of 2020/05/15, date of the 2^nd^ lockdown. b) Drop (%) in COVID-19 cases in different *comunas*.

## C Metapopulation Model with Age Structure

The model presented here is largely based on Ref. [36]. We consider *N* populations and *K* age groups. We introduce the matrix *C*^*j*^ ∈ ℝ^*K*×*K*^ whose element 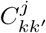 indicates the contacts rate between age group *k* and *k*′ in population *j*. Similarly, we introduce the matrix Σ ∈ ℝ^*N* ×*N*^, whose element *σ*_*ji*_ indicates the commuting rate from population *j* to *i*. We consider a *SLIR* compartmentalization setup with traveling Infectious, analogous to the case described in the main text (Tab. 1). In the following, we will use the notation 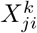 to indicate individual in age bracket *k* and compartment *X* living in population *j* and traveling to *i*. In these settings, we define the force of infection 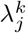 as the probability that a susceptible individual in age group *k* and in population *j* transit to the Latent compartment in the unit of time. This force of infection has two contributions, from the contacts occurred in the home population 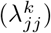 and in other connected populations 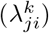. These can be written down as:

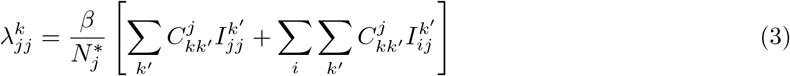

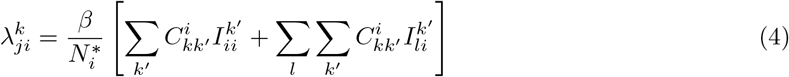

**Table 1:** Transitions between compartments

| Transition | Type | Rate |
| --- | --- | --- |
| $S_j^k \rightarrow L_j^k$ | Contagion | $\lambda_j^k$ |
| $L_j^k \rightarrow I_j^k$ | Spontaneous | $\epsilon$ |
| $I_j^k \rightarrow R_j^k$ | Spontaneous | $\mu$ |

Where *β* is the probability of becoming infected as a result of a single contact, and 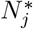 is the effective number of individuals in population *j* (more details below).

To use these expressions, we need to substitute the equilibrium values for 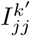 and 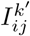. For a general compartment *X*, holds for consistency the following relation:

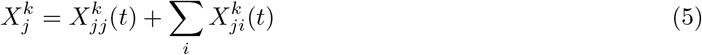

For 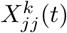 and 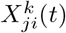 we can write the following rate equations:

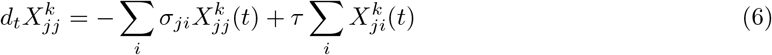

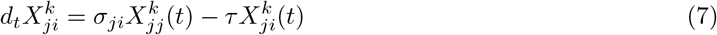

Where *τ* set the time scale of commuting (results in the main text are obtained for *τ* ^−1^ = 1/3 *day*). From Eq. 5 we derive 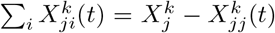. We substitute this expression in Eq. 6 and we define the total commuting rate of population *j* as *σ*_*j*_ = *Σ*_*i*_*σ*_*ji*_:

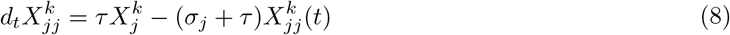

We solve Eq. 8 for 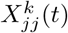:

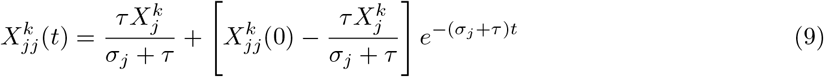

When *t* → ∞, we obtain the equilibrium value:

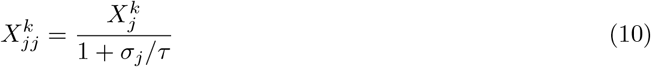

We take Eq. 7 and we impose the equilibrium condition 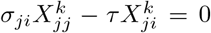. Then, substituting the result just derived, we obtain the equilibrium value also for 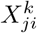:

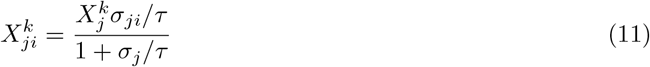

Therefore, substituting the equilibrium values in Eq. 3 and 4 we obtain:

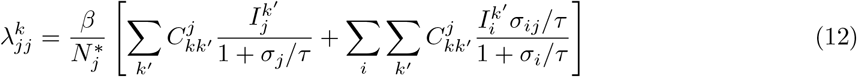

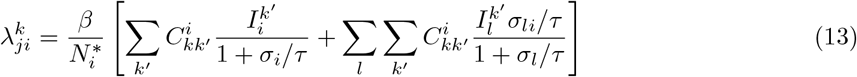

Then, the total force of infection 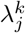 can be written as the the sum of these two contributions weighted, respectively, by the probability of finding a susceptible in age group *k* from population *j* in the home population 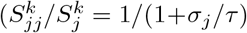 using Eq. 10), and in a connected population *i* (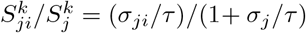 using Eq. 11):

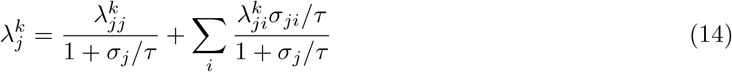

Finally, we still have to give an expression to 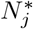, the effective number of individuals in population *j* introduced in Eq. 3. This effective population must consider both the individual from *j* present in *j* and those traveling to *j* from connected populations *i*. Using the equilibrium values derived in Eq. 10 and Eq. 11:

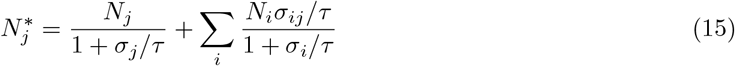

Mobility restrictions are implemented changing the commuting matrix Σ at the start of the partial and the full lockdown. Contacts reduction, instead, is implemented multiplying the contacts matrix *C*^*j*^ of each population (*comunas*) by the respective parameter. For example, the general element of the contacts matrix *C*^*j*^ during the partial lockdown for *comuna j* is 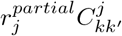.

